# Public attitudes towards COVID-19 contact tracing apps: A UK-based focus group study

**DOI:** 10.1101/2020.05.14.20102269

**Authors:** Simon N Williams, Christopher J. Armitage, Tova Tampe, Kimberly Dienes

## Abstract

**OBJECTIVE:** To explore public attitudes to the proposed COVID-19 contact tracing app in the United Kingdom.

**DESIGN:** Qualitative study consisting of five focus groups carried out between 1^st^-4^th^ May, 2020 (39–42 days after the official start of the UK lockdown).

**SETTING:** Online video-conferencing

**PARTICIPANTS:** 22 participants, all UK residents aged 18 years and older, representing a range of different genders, ages, ethnicities and locations.

**RESULTS:** Participants were split roughly equally in number across three groups: will use the app; will not be using the app; and undecided as to whether they will use the app. Analysis revealed five main themes: (1) Lack of information and misconceptions surrounding COVID-19 contact tracing apps; (2) concerns over privacy; (3) concerns over stigma; (4) concerns over uptake; and (5) contact tracing as the ‘greater good’. These themes were found across the sample and the three groups. However, concerns over privacy, uptake and stigma were particularly significant amongst those state they will not be using the app and the view that the app is for the “greater good” was particularly significant amongst those who stated they will be using the app. One of the most common misconceptions about the app was that it could allow users to specifically identify and map COVID-19 cases amongst their contacts and in their vicinity.

**CONCLUSIONS:** We offer four recommendations: (1) To offset the fact that many people may not be accessing, or might be avoiding, news coverage on COVID-19, authorities must communicate to the public via a range of methods including but not limited to: social media ads, postal information, text messaging and other emergency alert systems. (2) Communications should emphasise that the app cannot enable the user to identify which of their contacts has reported COVID-19 symptoms or tested positive. (3) Communication should emphasise collective responsibility (‘the greater good’) to promote social norms around use of the app (4) Communication should provide a slogan that maximises clarity of message, for example: ‘*Download the app, protect the NHS, save lives*’.

## Background

The current coronavirus (COVID-19) pandemic is widely considered the greatest challenge to public health in living memory. As of 7^th^ May 2020, COVID-19 accounted for a reported one-quarter of a million deaths globally, and 30,000 deaths in the UK alone, with total numbers of cases and deaths increasing daily [1]. The novelty of the SARS-CoV-2 virus means that no vaccines or antiviral drugs are available. In the absence of biomedical interventions to reduce morbidity and mortality from COVID-19, non-pharmaceutical interventions, such as physical distancing, hand hygiene and restricted movement have been proposed, including the “lockdown” policy of only leaving the home when essential (implemented in the UK on 23^rd^ March 2020) [2]. Although highly effective as a means to significantly slow the transmission of the virus (that is, to lower its reproductive (R0) value) in the population, lockdown is a very strict strategy that has a number of significant adverse impacts, including wider economic impacts for society [3] and social and psychological impacts for individuals [4]. As such, it is necessarily a temporary and relatively short-term measure.

One of the key components of many countries’ post-lockdown strategies is the use of contact tracing. Contact tracing is where those in close contact with individuals, who report symptoms indicative of an infectious disease and/or test positive for the disease, are identified and given appropriate medical instruction. It has successfully been used to control the spread of other novel communicable diseases like Ebola [5]. Traditionally, contact tracing relies on a team of health workers to track possible transmissions manually, through interviews with those who test positive for an infectious disease, in order to work out with whom they have recently been in close contact. Contacts deemed at risk are then informed of the fact they have been exposed and are then advised (or in some countries required) to self-isolate. Several countries, most notably South Korea and Singapore, have successfully used contact tracing (primarily using manual tracing) methods at an early stage in the outbreak in order to dramatically slow the spread of, and reduce the total number of cases and deaths from, COVID-19 [6].

However, whilst manual contact tracing has proven successful at early stages in the outbreak, where total number of cases are relatively low, it has been argued that doing so at a later stage (such as the current stage in the UK), when total numbers of infections are much higher, is problematic [7]. Firstly, manual contact tracing on a large scale is highly resource-intensive. For instance, it is proposed that in the UK, a contact tracing team of 18,000 people will be employed [8]. Secondly, it has been argued that due to the speed at which COVID-19 spreads, manual contact tracing might be too slow to be effective on its own [9]. As such digital contact tracing has been proposed and is being actively explored in a number of countries, including the UK, as a potential supplement to manual contact tracing [10].

On 4^th^ May 2020, the UK government revealed details of its app, developed by NHSX (the technology arm of the UK’s National Health Service), and began piloting the app on the Isle of Wight (a small island with a population of approximately 140,000) [11]. The app has attracted some criticism, as reported in the UK media, because of general concerns over privacy as well as because, unlike versions used or proposed in a number of other countries, it uses a ‘centralised model’ (i.e. where contact data is stored in a central database and not exclusively on the phones themselves) [12]. Although at time of writing, subsequent plans for the full release of the app to the total UK population were yet to be confirmed, it was suggested that the app would be rolled out sometime in May 2020 [13]. Modelling evidence suggests that in order for the app to be effective, use and adherence would need to be very high, with 80% of smartphone users (equivalent to 56% of the UK population) needing to use the app in order to completely stop the pandemic spreading, although lower rates of use could still have beneficial impacts [9].

Research is beginning to explore public attitudes towards potential exit strategies, although much of it so far has taken the form of public opinion surveys [14] [15]. Some surveys have suggested that a majority of smartphone users would use the app [16], but emerging evidence from early adopting countries like Singapore suggests that actual uptake might be considerably lower [17]. There is, to our knowledge, as yet no published qualitative research on public attitudes towards COVID-19 contact tracing apps. Qualitative research can provide an important supplement to large sample opinion polls involving an in-depth exploration of the reasons and motivations behind individuals’ intentions towards use of contact tracing apps. If the UK’s contact tracing strategy is to be effective, government and health authorities will need to understand and respond quickly to any public concerns over the app. The aim of this paper is to explore attitudes towards the proposed contact tracing app in a diverse sample of the UK public. Specifically, we seek to explore people’s knowledge of the contact tracing app, their views on the app (specifically the extent to which they are favourable to it or not), and ultimately whether they plan to use the app or not. These findings can feed into debates around how best to communicate to the public the importance of contact tracing as a means to reduce the spread of, and prevent further waves of, the virus, which in turn could prevent the need for further lockdowns.

## Methods

We conducted five online focus groups with 22 participants between 1^st^ and 4^th^ May 2020 during the UK’s COVID-19 lockdown (39–42 days after the official start of the UK lockdown). Participants were adults aged 18 years or over currently residing in the UK. Online focus groups were methodologically necessary due to the fact that in-person focus groups were not feasible due to the pandemic. However, online focus groups do have methodological benefits particularly as a cost-effective method of eliciting public views from a geographically dispersed group of participants [18][19].

Focus groups were conducted as part of a larger, mixed-methods longitudinal study tracking public attitudes to COVID-19 and its associated policies in the UK, as well as the social and psychological impacts of such policies. Focus group recruitment took place online through a combination of social media snowball sampling, online community and volunteer advertising sites and social media advertisement. We took a purposive approach to sampling, attempting to include a range of ages, genders, racial/ethnic identities and occupational backgrounds (Table 1).

**Table 1:** Demographic details reported by participants.

| Characteristic | N (%) |
| --- | --- |
| *Gender* |  |
| Female | 10 (45) |
| Male | 12 (55) |
| *Age range* |  |
| 18-29 | 8 (36) |
| 30-39 | 3 (14) |
| 40-49 | 6 (27) |
| 50+ | 1 (5) |
| Undisclosed | 4 (18) |
| *Ethnicity* |  |
| White - British | 13 (59) |
| White – any other White background | 2 (9) |
| Asian or Asian British - Pakistani | 4 (18) |
| Undisclosed | 3 (14) |

Focus groups met virtually via the web video-conferencing platform Zoom for approximately one hour each. Most participants joined using both video and audio, although some were given the option, and preferred, to join with audio only. All focus groups were organised and moderated by SW (a medical social scientist). The topic guide for the focus groups was initially developed by the research team in virtual group meetings. Although the focus groups discussed topics related to the broader research project (e.g. experiences on social distancing, views on COVID-19 testing and vaccinations), it also specifically included questions seeking to elicit participants views on covid-19 contact tracing apps (e.g. ‘do you think you will use the planned covid-19 app?’ and ‘what are your views on the planned covid-19 contact tracing app?’

### Analysis

Data were collected and analysed in accordance with a Grounded Theory approach [20]. This is an iterative process through which emergent themes from early focus groups were used to add to or refine questions and prompts during subsequent groups. Data were transcribed for coding, with all participant information being anonymised to protect confidentiality. SW and KD analysed the transcripts and developed and applied the thematic coding framework, moving from open through to more focused coding [20][21]. Themes were discussed and developed with CJA and TT during virtual research group meetings. Negative case analysis was used to seek for information that did not fit emergent themes, and where this occurred, themes were modified accordingly [22]. Data collection and analysis continued until theoretical saturation was reached [21]. Data were analysed in NVivo (version 11.4.3, QRS).

## Results

Participants fell into three groups regarding whether or not they felt they would use the contact tracing app. One group of participants stated they will be downloading the contact tracing app (‘would use app’) (n = 7), one group stated they will not (‘will not use app’) (n = 7), and a third group were undecided (‘undecided’) (n = 8). No patterns emerged according to the demographics of the participants, with a mix of genders, ages and occupational groups across all three groups.

Analysis revealed five main themes: (1) Lack of information and misconceptions surrounding Covid-19 contact tracing apps; (2) concerns over privacy; (3) concerns over stigma; (4) concerns over uptake; and (5) contact tracing as the ‘greater good’. These themes were found across the sample and the three groups, however certain themes were more apparent, and appeared to carry more weight in influencing their views as to whether they would use the app or not, in some groups compared to others. Many participants across all three groups either expressed a lack of knowledge of, or displayed misconceptions about, COVID-19 contact tracing apps. Concerns over privacy and uptake were expressed across all three groups, but seemed to be particularly significant for those who said they would not use the app. Concerns over stigma were expressed by participants in the ‘would not use app’ and ‘undecided’ groups, but not in the ‘would use’ group. The view that contact tracing was for ‘the greater good’ was expressed by participants in the ‘would use’ and ‘undecided’ groups, but not in the ‘would not use’ group, and was particularly significant for those in the ‘would use’ group.

### Lack of information and misconceptions surrounding Covid-19 contact tracing apps

Many participants either expressed a lack of knowledge of contact tracing and contact tracing apps or appeared to have misconceptions over what the official UK government-proposed app entails. Amongst those who had not heard of contact tracing apps, one of the reasons was because they had “*not been watching the news*” (Participant 19, female, 20s), as they were “*sick and tired of hearing more of coronavirus*” (Participant 20, female, 20s). Amongst those who had heard of them, one of the biggest misconception was that the app would entail some form of ‘mapping’ that would be visible to other smartphone users or the public in general:

> “*I think they wanted to develop an app or something, so you would know how many Covid cases are in your area, so you know where not to go. So, if everyone got sent a test and managed in certain areas not go out until everyone has been tested, so you know where the virus is so you know who can leave their homes and who can’t*” (Participant 6, male, 20s)

### Concerns over privacy

The most commonly stated concern was over data privacy and security. Participants expressed a reluctance to have their data accessed by government or health authorities. Participants associated contact tracing with increased surveillance by government, and were concerned by what they perceived as “submitting” their personal information:

> “*Contact tracing seems quite Big Brotherly. I don’t think I am willing to submit all my data and all of my contacts for the government to scrutinise who I see regularly. I don’t think I will be willing to join the contact tracing apps*” (Participant 16, female, 30s)

Participants were also concerned that their data would be accessible to others outside of government and health authorities, including “third parties” or “hackers”:

> “*You don’t know who is running these apps, you don’t want third party accessing your information. This is the worrying thing about modern technology; you do not know who is accessing your health and things*.” (Participant 10, male, 40s)
>
> *“Hackers could pose as health experts or something, there are so many scams that are going on. Scammers are supposedly selling. I am not in favour of this app personally.”* (Participant 7, male, 20s)

However, some of these concerns over privacy seemed to stem from the misunderstanding discussed in the previous section that app users would be able to identify specific covid-19 cases amongst their contacts or their vicinity:

> “*Imagine walking around, and thinking ‘you’ve got it, you’ve got it’ and you have got it [phone] in your hand … Imagine you were like, oh ill go see my friends when we are all allowed, and they were like ‘You have got corona it says it on my app’”* (Participant 19, female, 20s)

For those who stated they will not use the app, privacy was the most common reason stated for their decision. They were generally opposed to anyone, including government and health officials accessing their data under any circumstance. For those who said they would download the app, privacy concerns were mentioned, but were seen to be outweighed by the perceived benefits of the app for public health (as discussed below). Those who were undecided tended to argue that they would use the app provided they were sure that their data was not visible to other smartphone users or the general public they were given assurances over privacy and data protection:

> “*I would get it if there were no privacy concerns and it was just for the government to track the spread of the virus … but I wouldn’t do it if there was a map to see*.” (Participant 18, female, 20s)

Those who were undecided also tended to weigh up their concerns over data privacy with their view that apps which require the disclosure of personal information are already commonplace:

> “*I’m a bit waryy with the app but I tend to download anything and sign my life away all the time … I think I would be alright with that kind of stuff …. I think everyone is already traced and they don’t even know*.” (Participant 16, female, 30s)

### Concerns over stigma

Another commonly expressed concern was over the stigmatizing potential of the app. This concern was related to concerns over a lack of privacy and specifically the misconception that the app would enable people to use the app to identify others that have or have had covid-19:

> “*I actually think that [the contact tracing app] is a terrifying concept… it’s like being branded with a. horrendous black mark. … I could look and be like my friend, my neighbour has got Covid.”* (Participant 17, male, 20s)

These participants feared that being able to identify others with covid-19 would lead to discrimination:

> “*It could cause hate crime as well. Finding out ‘oh you know I got it from this person’, and finding out your neighbours have it, and ‘oh I will stay away from them’*.” (Participant 9, female, 30s)
>
> “*It is just asking for more isolation and hysteria. … It would be like Snap Maps: ‘oh I want to go to Tesco, but oh no she’s there and she has got Covid’…. I think that would make me not want to get tested [for covid-19]. I wouldn’t want to be on the ‘Snapmap of Shame’. Like, if you could see all these people that you know have corona, that could lead to bullying in some cases.”* (Participant 19, female, 20s)

### Concerns over uptake

Another commonly voiced concern was over uptake of the app. A number of participants questioned whether enough people would use it in order for it to be an effective means of reducing the spread of the virus:

> “*We have only reduced or flattened the curve because we have all collectively socially distanced … and if people were to download the app I think it would have to be done in a large group of people if not everybody for it to be effective … I don’t know how effective it would be in terms of transmission rates. I wouldn’t rush to download it*.” (Participant 17, male, 20s)

Amongst those who had heard of contact tracing, some drew comparisons between the UK and countries who had experienced the pandemic earlier and who had already made, in their view, effective use of contact tracing, such as Singapore. However, these participants tended to argue that the high level of adherence and enforcement in such countries would not be achievable in the UK which was perceived as less accepting of state interventionism and less collectivist:

> *“The issue here is will society take it on board and actually do it. I think one of the reasons why places like China, South Korea, Singapore, those Asian countries can successfully manage those situations is because they have a relatively compliant society, people tend to work together or maybe it’s just because they are used to having their civil liberties curbed to a degree”* (Participant 12, male, 40s)

Another consideration was that a proportion of the population did not have smartphones, for example the homeless, elderly or socially or economically vulnerable in society. For some, such inequalities in access to the technology would reduce the effectiveness of using an app as part of a contact tracing strategy:

> “*Some people don’t know how to download an app, some people don’t have a smartphone, some people may be left out, the vulnerable, and if they aren’t able to put in the data this might have an effect on other people. So how accurate and how up to date and how reliable the information could be would make a difference*” (Participant 1, male, 30s)

### Contact tracing as “the greater good’

No participants were unequivocally positive about the contact tracing app. Ultimately, what distinguished those who were intended to use the app from those who did not intend on using it, was their belief that it was the ‘right thing’ to do because it would be beneficial for the wider public health, and that this potential benefit outweighed their concerns:

> *“If it is going to be for the greater good, I would do it, but it doesn’t fill me with joy.” (*Participant 17, male, 20s)
>
> *“I would do it if it helps but it’s not something I want to do.”* (Participant 8, female, 40s)

In particular the recognition of the severity and urgency of the pandemic was seen as a reason as a reason to use the app:

> *“I would really support it, I know privacy is really important … anything that would help, it doesn’t make sense why people wouldn’t participate; people are dying all over the world, what’s more important at the moment, to try and stop this or people knowing what’s happened on your phone*” (Participant 16, female, 30s)

These participants implied that, because contact tracing in their view was necessary for slowing the spread of the virus, they had little agency over whether or not to use the app. Contact tracing was here characterized as an unavoidable exit strategy, as the “only way out”:.

> “*The idea of tracking, with a Big Brother or something, I am totally opposed to it normally, but I can’t tell you how totally opposed normally. But based on what countries that have been successful have done, I feel like this might be the only way out of this*.” (Participant 21, female, 50s)

## Discussion

Our findings reveal that the UK public are split over whether they feel they will use the proposed COVID-19 contact tracing app. We found three, roughly equally sized, groups: those who feel they will download the app, those who won’t and those who are undecided. A major theme was the perceived lack of knowledge and the existence of misconceptions around the app. One of the common misconceptions was that the app would provide identifiable information, and users themselves would be able to specifically identify others with COVID-19 (or be identified themselves). Three main concerns were raised: concerns over privacy; concerns over stigma; and concerns over uptake. These concerns were expressed by participants across all three groups, but, as discussed below, were more salient in some groups over others, in relation to influencing their view on whether they will likely use the app or not.

The fact that the groups were roughly equal in proportion contrasts with recent surveys that suggest a very high (∼80%) proportion of the UK public will either definitely or probably install the app [16]. It is of course a limitation of this study that its sample size, which is small in relation to such large quantitative surveys, does not permit our findings to be readily generalized to the UK population as a whole, and so the findings on the proportions of who plans to use the app or not should be treated cautiously and followed up with further research. This however is a limitation inherent to all qualitative research and not specifically to the current study. The contribution of this study is its ability to shed light on the underlying reasons and beliefs that account for people’s views on the app and which ultimately shape their decision of whether or not to use it.

Concerns over stigma stemmed from a fundamental misconception that the app could enable its users to identify COVID-19 cases amongst their contacts or even synchronously map cases near them. It is worth noting that, despite these misconceptions, such findings imply that COVID-19 may be, for some, becoming a stigmatised disease. Future research will explore the potential stigmatization of COVID-19 sufferers in more depth and over time. The main implication of relevance to the present study is that the public may not be adequately informed as to what the app entails. Improved communication regarding the purpose and nature of the app will likely lead to increased use of the app (see implications section below). Concerns over uptake were framed in terms of social inequalities and cultural norms around state interventionism. It was argued that because certain social groups do not have access to smartphones, and because the UK public would not be willing to accept contact tracing in general as a strategy, it would not be as effective as it had been in countries like Singapore or South Korea. Of course, the risk here, from a public health perspective is that if enough people feel that it is not going to be effective because not enough people are going to use it (and use this as a reason to not use it themselves) then their beliefs may contribute to a self-fulfilling prophesy at the population level. Thus, normative beliefs in the potential effectiveness of the app amongst the UK public are necessary in order to maximise uptake (see implication section below). Concerns over privacy were often framed in terms of a general lack of trust or faith in government and its handling of the pandemic thus far, and their opposition to what was perceived as an unnecessary form of government control and the role of technology in the rise of a surveillance society [23][24][25].

Decision-making was heavily influenced by moral reasoning. Those who said they would not download the app were motivated primarily by their moral opposition to the use of such technologies as an infringement of individual civil liberties. By contrast, among those who were intending on using the app, decision-making was driven by a more utilitarian evaluation of the relative costs and benefits. Many shared the same concerns as those who did not intend to use the app, but deemed that the potential life lost from COVID-19 outweighed any privacy infringements. Arguably, these participants viewed using the app to be a decision high in ‘moral intensity’, because of the temporal immediacy of the pandemic (its rapid spread), the magnitude of its consequences (its high death rates), and its concentration of effect (i.e. severe impacts (serious illness and death) amongst a relatively small number of people being greater than modest impacts (privacy risks) amongst a much larger number of people (total app users)) [26]. Finally, those who were undecided were generally still in the process of a utilitarian evaluation of the perceived costs and benefits of the contact tracing app. Therefore, if the UK is to achieve the high uptake levels that would be required in order to stop or even substantially reduce the spread of the virus, it is necessary to achieve the support of those who are as yet undecided on whether they are likely to use the app. Some ways in which governments and public health authorities can better communicate to the public around the COVID-19 contact tracing app are discussed below.

### Limitations

As discussed above, one of the limitations of this study is, like all qualitative research, it is not possible to generalize findings (for example on the proportions of how many people plan to use the app) to the wider UK public. However, our qualitative research will also be used to inform future quantitative research exploring patterns and factors related to intentions and beliefs concerning Covid-19 contact tracing apps. For example, this research can inform the development of survey questions related to underlying beliefs concerning the nature of the app, normative beliefs concerning protecting the vulnerable (the ‘greater good’), and concerns over privacy, uptake and stigma.

This study is also limited insofar as it did not recruit participants from ‘clinically extremely vulnerable’ categories [27]. Additionally, the study did not include any individuals aged 70 or older (also considered ‘clinically vulnerable’, although this was because we did not receive any responses from this age group, despite noting in our recruitment material that applications from those considered clinically vulnerable were particularly encouraged. Future research will specifically explore the attitudes of those clinically extremely vulnerable and clinically vulnerable on COVID-19-related policy.

### Recommendations for policy and practice

Based on our findings, we suggest the following recommendations for policy and practice concerning engaging and communicating with the public regarding the proposed COVID-19 contact tracing app:

1. Government and public health authorities need to consider a range of methods to engage and communicate to the public as to the purpose and nature of the contact tracing apps. Although traditional media play an important role in disseminating information, our findings suggest that there is a proportion of the population who may be actively avoiding news coverage on coronavirus (as a result of over-exposure to Covid-19 coverage and/or as a coping or avoidance strategy). Indeed, limiting watching news coverage on COVID-19 has been recommended as a means to protect mental health during the pandemic [28], and some people may be significantly avoiding news coverage. As such, authorities must explore a range of methods and media to communicate the purpose and nature of contact tracing apps, including but not limited to the use of: social media ads, postal information, text messaging and other emergency alert systems.
2. Communications should succinctly explain that the app cannot enable the user to identify which of their contacts has reported COVID-19 symptoms or tested positive, and that confidentiality is protected. The UK government should reconsider the decision to adopt a decentralised system. Concerns over “Big Brother” government surveillance will likely be allayed for many by a switch to a decentralised system. This could in turn have a positive effect on app uptake and use.
3. Government and public health authorities need to continue to harness the message that contact tracing is a measure that can contribute to “the greater good” in terms of saving lives and reducing the spread of the disease. Amongst those who intend to use the app, this was the key factor driving their decision. Linking to the ‘moral intensity’ of Covid-19 pandemic, specifically its high death rates and the potential threat of a second wave is also likely to be effective in facilitating uptake of the app. Similar to the campaign over ‘stay at home, protect the NHS, save lives’ campaign for lockdown, the proposed app should provide a slogan that maximises clarity of message, for example: ‘*Download the app, protect the NHS, save lives*’.
4. In order to maximise uptake, normative beliefs will need to reflect the potential effectiveness and popularity of the app. In this way, public health messages could be similar to those employed in the effort to encourage widespread adherence to social distancing guidelines, including campaigns around collective responsibility (‘stick together by staying apart’).

## Data Availability

Ethical restrictions related to participant confidentiality prohibit the authors from making the data set publicly available. During the consent process, participants were explicitly guaranteed that the data would only be seen my members of the study team. For any discussions about the data set please contact the corresponding author, Simon Williams.

## Declarations

### Transparency declaration

The lead author (the manuscript’s guarantor) affirms that the manuscript is an honest, accurate, and transparent account of the study being reported; that no important aspects of the study have been omitted; and that any discrepancies from the study as planned (and, if relevant, registered) have been explained.

### Authors’ contributions

All authors contributed to the planning of the study. The analysis was conducted by SW and KD. The initial draft of the article was written by SW. All authors revised the manuscript and approved the final version for publication. SW is the guarantor of the article.

### Ethics statement

Ethical approval was received by Swansea University’s Research Ethics Committee.

## References

1. European Centre for Disease Prevention and Control. COVID-19 situation update worldwide, as of 7^th^ May 2020. Available at: https://www.ecdc.europa.eu/en/geographical-distribution-2019-ncov-cases (Accessed 7th May 2020)

2. National Health Service (NHS) UK. Advice for everyone: Coronavirus (COVID-19). Available at: https://www.nhs.uk/conditions/coronavirus-covid-19/ (Accessed 14th May 2020).

3. Fernandes N. Economic impacts of coronavirus outbreak (COVID-19) on the word economy. Social Science Research Network. Available at: https://papers.ssrn.com/sol3/papers.cfm?abstract_id=3557504 (Accessed 14th May 2020).

4. Williams S, Armitage C, Tampe T, Dienes K. Public perceptions and experiences of social distancing and social isolation during the COVID-19 pandemic: A UK-based focus group study. medRxiv 2020.04.10.20061267; doi: https://doi.org/10.1101/2020.04.10.20061267

5. World Health Organization. Contact Tracing (website). Available at: https://www.who.int/csr/disease/ebola/training/contact-tracing/en/ (Accessed 14th May 2020)

6. Wong J, Leo Y, Tan C. COVID-19 in Singapore: Current Experience. JAMA 2020; 323:1243–1244.

7. Keeling M, Hollingsworth D, Read J. The Efficacy of Contact Tracing for the Containment of the 2019 Novel Coronavirus (COVID-19) MedRxiv 2020. Available at: https://www.medrxiv.org/content/medrxiv/early/2020/02/17/2020.02.14.20023036.full.pdf

8. Vaughan A. UK sets new target to recruit 18,000 contact tracers by mid-May. New Scientist 28 Apr 2020. Available at: https://www.newscientist.com/article/2242088-uk-sets-new-target-to-recruit-18000-contact-tracers-by-mid-may/ (Accessed 14 May 2020)

9. Ferretti L, Wymant C, Kendall M, Zhao L, Nurtay A et al. Quantifying SARS-CoV-2 transmission suggests epidemic control with digital contact tracing. Science 368: eabb6936.

10. Public Health England. Letter to Directors of Public Health about contact tracing (1st May) 2020. Available at: https://assets.publishing.service.gov.uk/government/uploads/system/uploads/attachment_data/file/882950/RGleave_letter_to_DSPH2.pdf (Accessed 14th May 2020).

11. Department of Health and Social Care, UK. Coronavirus test, track and trace plan launched on Isle of Wight. Available at: https://www.gov.uk/government/news/coronavirus-test-track-and-trace-plan-launched-on-isle-of-wight (Accessed 14th May 2020).

12. Hern A, Proctor K. UK may ditch NHS contact-tracing app for Apple and Google model. The Guardian. 7th May 2020. Available at: https://www.theguardian.com/technology/2020/may/07/uk-may-ditch-nhs-contact-tracing-app-for-apple-and-google-model (Accessed 14th May 2020).

13. National Cyber Security Centre. NHS COVID-19: the new contact-tracing app from the NHS. Available at: https://www.ncsc.gov.uk/information/nhs-covid-19-app-explainer. Accessed 14th May 2020.

14. Geldsetzer P. Knowledge and perceptions of COVID-19 among the general public in the United States and the United Kingdom: A cross-sectional online survey. Annal Intern Med 2020; E-pub ahead of print, DOI: 10.7326/M20-0912.

15. Altmann S, Milsom L, Zillessen H, Blasone R, Gerdon F et al. Acceptability of app-based contact tracing for COVID-19: Cross-country survey evidence. MedRxiv 2020; Available at: https://doi.org/10.1101/2020.05.05.20091587

16. Milsom L, Abeler J, Altmann S, Toussaert S, Zillessen H et al. Survey of acceptability of app-based contact tracing in the UK, US, France, Germany and Italy 2020. Available at: https://osf.io/7vgq9/ (Accessed 14th May 2020).

17. Vaughan A. There are many reasons why covid-19 contact-tracing apps may not work. New Scientist. Available at: https://www.newscientist.com/article/2241041-there-are-many-reasons-why-covid-19-contact-tracing-apps-may-not-work/ (Accessed 14th May 2020)

18. Tates K, Zwaanswijk M, Otten R, et al. Online focus groups as a tool to collect data from hard-to-include populations: examples from pediatric oncology BMC Med Res Method 2009;9:ar15.

19. Williams S. A 21^st^ century Citizens’ POLIS: Introducing a democratic experiment in electronic citizen participation in science and technology decision-making,’ Public Understanding of Science 2010;19:528–544.

20. Coffey A, Atkinson. Making Sense of Qualitative Data. London: Sage. 1996

21. Silverman D (Ed.). Qualitative research: theory, method and practice. London: Sage, 1997.

22. Mays N, Pope C. Rigor and qualitative research. BMJ 1995;311:109–112.

23. Foucault M. Discipline and punish: The Birth of the Prison 1977; New York: Random House, 1977.

24. Lyon D. Surveillance society: Monitoring everyday life 2001; Buckingham: Open University Press.

25. Lupon D. M-health and health promotion: The digital cyborg and surveillance society. Social Theory & Health 2012; 10: 229–244.

26. Jones T. Ethical decision making by individuals in organizations: An issue-contingent model. The Academy of Management Review 1991; 16: 336–395.

27. NHS UK. People at higher risk from coronavirus (website) 2020. Available at: https://www.nhs.uk/conditions/coronavirus-covid-19/people-at-higher-risk-from-coronavirus/. Accessed: 14th May 2020.

28. NHS UK. Mental wellbeing while staying at home. Available at: https://www.nhs.uk/oneyou/every-mind-matters/coronavirus-covid-19-staying-at-home-tips/. Accessed 14th May 2020.

